# Beyond skills: The impact of personality traits, empathy and stress mindset on OSCE outcomes

**DOI:** 10.64898/2026.04.14.26350876

**Authors:** Douglas Henderson, Baptiste Lignier, Bernard Moxham, Odile Plaisant, Université Paris Cité OSCEs study group, Céline Buffel du Vaure, Albert Faye, Donia Bouzid, Cédric Lemogne, Alexis Guédon

## Abstract

Objective Structured Clinical Examinations (OSCEs) are widely used to assess medical students’ clinical skills, including non-technical abilities such as communication and empathy. However, the potential influence of individual psychological traits—such as personality dimensions, empathy, and stress-related mindset—on OSCE performance remains understudied. This study investigated associations between personality traits, empathy levels, stress mindsets, and performance in OSCEs among medical students.

An online questionnaire (including the Big Five Personality Traits Inventory 2, the Jefferson Scale of Physician Empathy (Medical Student version), the Growth Mindset Scale, the Stress Mindset Measure) was provided to all fifth-year medical students enrolled at the Université Paris Cité for six weeks before undertaking graduation summative OSCEs. Their scores were correlated with OSCE performance using Spearman’s correlation and linear regression analyses.

A total of 99 questionnaires were included and analysed. None of the psychometric tests we assessed showed a significant correlation with OSCE scores. The strongest predictors of success in OSCEs were higher scores in written examinations, previous OSCE performance, and being female. In non-interactive OSCE stations, conscientiousness was the only significant predictor, with a positive association (p=0.001). Neuroticism was positively associated with performance improvement between OSCE sessions (p=0.042).

Personality traits, self-reported empathy, and stress-related mindsets do not predict success in OSCEs as isolated traits. Further research is needed to determine whether it holds true for all kinds of OSCEs. Multidimensional psychometric assessment may be relevant when investigating performance outcomes in OSCEs.

## Introduction

Objective Structured Clinical Examinations (OSCEs) have become the benchmark for assessing the clinical skills of medical students, including non-cognitive abilities [1–4]. Much research has tried to identify predictive factors for academic achievement in OSCEs, psychological features such as personality traits, anxiety, stress, or empathy [5–16] being investigated. These studies aim to adapt students’ training and preparation for OSCEs.

The Big Five Personality Inventory [17,18] that assesses extraversion (E), agreeableness (A), conscientiousness (C), emotional instability or neuroticism (N), and openness or open-mindedness (O) has been the most studied personality model [19]. Its ability to predict healthcare students’ academic achievements has been explored extensively. A recent systematic literature review indicates that conscientiousness is linked to higher academic performance in the context of health professionals’ training curricula [20]. Another study suggests that conscientiousness may enhance learning during preclinical years but not during clinical years [21]. Other studies however find no correlation [5,7].

Empathy encompasses some critical clinical skills, such as accurately perceiving patients’ emotions and effectively communicating physicians’ understanding, and the Jefferson Scale of Physician Empathy (JSPE) is a widely used assessment tool for this muti-dimensional construct in the context of medical care [22]. The JSE authors found a correlation between clinical competence and the JSPE [23].

Mindset theories are a fast-growing interest in the education of healthcare professionals [24–34]. As the authors define it, a mindset is “a mental frame or lens that selectively organizes and encodes information, thereby orienting an individual toward a unique way of understanding an experience and guiding one toward corresponding actions and responses [35]”. It has recently been suggested that a person’s state of mind can modulate their response to stress [36]. Recent reports indicate that a synergistic stress-related mindset intervention (namely “stress-can-be-enhancing mindset” and “growth mindset”) in high school students can transform stress perception and might enhance academic achievement [37].

We prospectively followed a cohort of fifth-year medical students, assessing several non-cognitive markers to identify independent predictors of academic achievement in OSCEs. We hypothesized that higher OSCE scores are achieved by students with higher agreeableness (A) and conscientiousness (C) personality characteristics, greater empathy, and stress-resilience.

## Methods

### Participants and setting

Eligible participants at the Université Paris Cité for this study were fifth-year medical students who had given their consent, had completed an online questionnaire between April 22nd, 2024 and June 4th, 2024, and had taken the OSCEs on June 5th, 2024. There were no exclusion criteria. All predictors were assessed in the six weeks immediately prior to the OSCEs. Finally, from July 15th to 17th 2024, fifth-year students also participated in a large planned written mock examination involving several medical schools. Written examination scores were assessed on a single comprehensive examination that covered all medical knowledge required for entering residency. Furthermore, OSCE stations could be dichotomised into interactive stations (i.e., involving a patient or a healthcare professional) and non-interactive stations (i.e., procedural or without human interaction). Progress in OSCE performance was assessed by comparing the results with those obtained in previous OSCEs (December 13^th^, 2023)

### Structured questionnaire

This study’s questionnaire was self-administered through an online platform (LimeSurvey GmbH, Hamburg, Germany) and consisted of 102 items (**Supplementary material**). It employed the following psychometric tests: the Big Five Inventory 2 (BFI2-Fr) [38], the Jefferson Scale of Empathy – Medical Students version (JSPE-MS)[39,40], the Growth Mindset Scale (GMS) [41,42] and the Stress Mindset Measure (SMM) [25]. Additionally, students were asked to self-report their gender, age, specialty choice, parenthood, native language, parents’ native language, parental education level, previous stage exposure (performing in public in an artistic or other field), and aspects of their clinical rotations (motivation and involvement, negative experiences, acquisition of clinical skills).

### Consent

All participants gave their informed consent. All required authorisations were obtained from the *CNIL* (National Committee for Information Technology and Civil Rights), and the local ethics committee (Institutional Review Board of the Université Paris Cité, Paris, France, number 00012024-21). The LimeSurvey GmbH platform complies with the General Data Protection Regulation (GDPR) of the European Union.

### Personality assessment

BFI-2-Fr [38,43] sixty items are scored on a five-point Likert scale ranging from one to five. E, A, C, N, O domains are measured by twelve items each. Fifteen facets are also assessed by four items each (Sociability, Assertiveness, Energy Level, Compassion, Respectfulness, Trust, Organization, Productiveness, Responsibility, Anxiety, Depression, Emotional Volatility, Intellectual Curiosity, Aesthetic Sensitivity, and Creative Imagination).

### Empathy assessment

The JSPE-MS [40] is a self-administered questionnaire that is comprised of twenty items and that uses a seven-point Likert scale. It explores three dimensions: “Perspective Taking”, “Compassionate Care” and “Walking in Patient’s Shoes”[44].

### Stress-related Mindsets assessment

In accordance with the idea of a synergistic interaction between Growth and Stress-can-be-enhancing Mindsets [37], we used two different scales. The SMM is an eight-item questionnaire, using a five-point Likert scale [36]. It determines whether a subject believes that stress can be beneficial or harmful. The GMS consists of a three-item questionnaire that uses a six-point Likert scale [41,42]. It determines a subject’s view of their own intelligence/cognitive ability, considering it to be either a fixed trait or a quality that can be developed.

### Comparison to other cohorts

Our study cohort was compared with the overall cohort of fifth-year medical students enrolled at the Université Paris Cité, **Supplementary material**.

### Statistical analysis

Primary analysis: to evaluate whether psychometric data could predict OSCE scores, we conducted Spearman correlation tests and performed linear regression analyses, including both univariate and multivariate models.

Secondary analyses: we examined the relationship between OSCE results and other data collected in the questionnaire, and investigated associations between psychometric tests and written exam results.

Calculations were made with R (R Core Team, 4.4.1), Rstudio (2023.06.1+524). Quantitative variables were described by their median and interquartile range unless otherwise stated. Symmetric associations between quantitative variables were described by Spearman correlations. Associations between qualitative variables were explored by Fisher exact tests.

Associations between explanatory qualitative/quantitative variables (psychometric tests, demographic data, other examination results) and a quantitative response variable: the beta coefficients (and their 95% confidence intervals) of univariate and multivariate linear models without regularization were provided using the *finalfit* package.

Associations between explanatory quantitative variables and a qualitative response variable: when testing whether groups differ in the distribution, a given quantitative variable either Wilcoxon rank sum tests (two-group comparisons) or Kruskal-Wallis (more than two groups) were used. Although the scores of the OSCE stations comprised items assessed either binarily or using a Likert scale, the final score was expressed out of 20 and was therefore treated as a continuous variable.

The questionnaire’s reliability over two consecutive trials was evaluated with a two-way agreement intraclass correlation coefficient following Koo and Li guidelines for selection and interpretation [45].

Heatmap representations relied on the *pheatmap* package. Results with p<0.05 were considered statistically significant. Adjustment for multiplicity of testing were done at the table level using the Benjamini-Hochberg procedure.

A *post hoc* exploratory cluster analysis was performed. Continuous variables including personality traits, the three dimensions of the JSPE, the SMM, and the GMS were centered and scaled, then reduced to four principal components (Kaiser criterion). K-means clustering (elbow method) was subsequently applied to the PCA scores, and cluster stability was assessed using bootstrap methods.

## Results

Among the 830 fifth-year medical students of the overall cohort, 103 completed at least one psychometric test on the online platform. Four of them did not take part in the OSCEs. We finally included 99 surveys. All participants completed the BFI-2-Fr, 89/99 (90%) completed the JSPE-MS, 84/99 (85%) completed the GMS and the SMM, and 83/99 (84%) responded to the demographic and clinical rotations questions. Among the 99 students included in our study, 7/99 (7%) did not take part in the written mock examination. 60/99 (61%) were female students, with a mean age of 23 years old (SD 3.74) (missing data 16%), **Table 1**. Compared to the entire fifth-year medical students cohort, participants had better results in previous OSCEs (p=0.009) and the written mock examination (p=0.010) but not in the OSCEs. The study cohort was similar in gender but not in age (p=0.008) to the overall cohort of fifth-year medical students, **Supplementary material**.

**Table 1:**
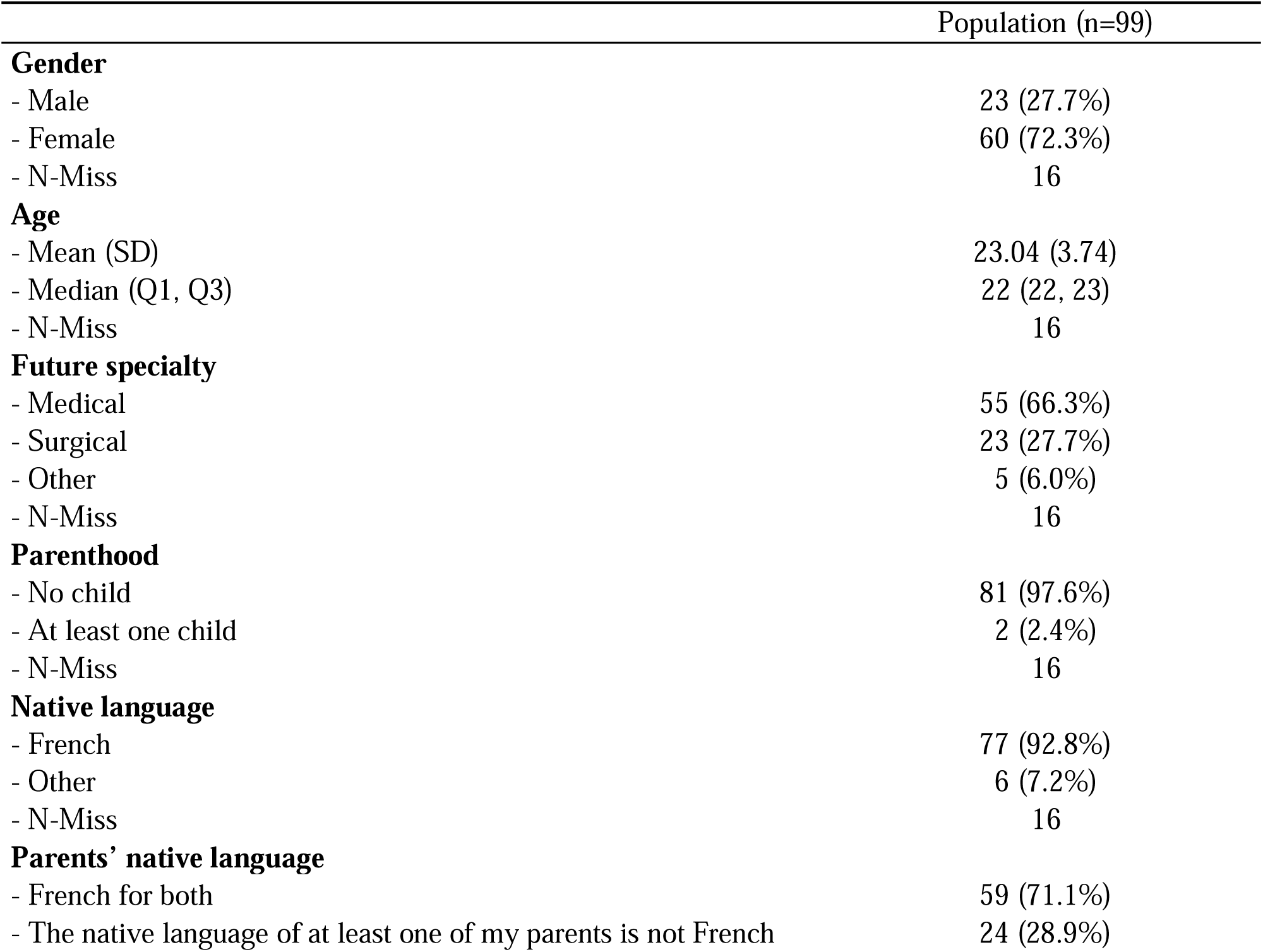
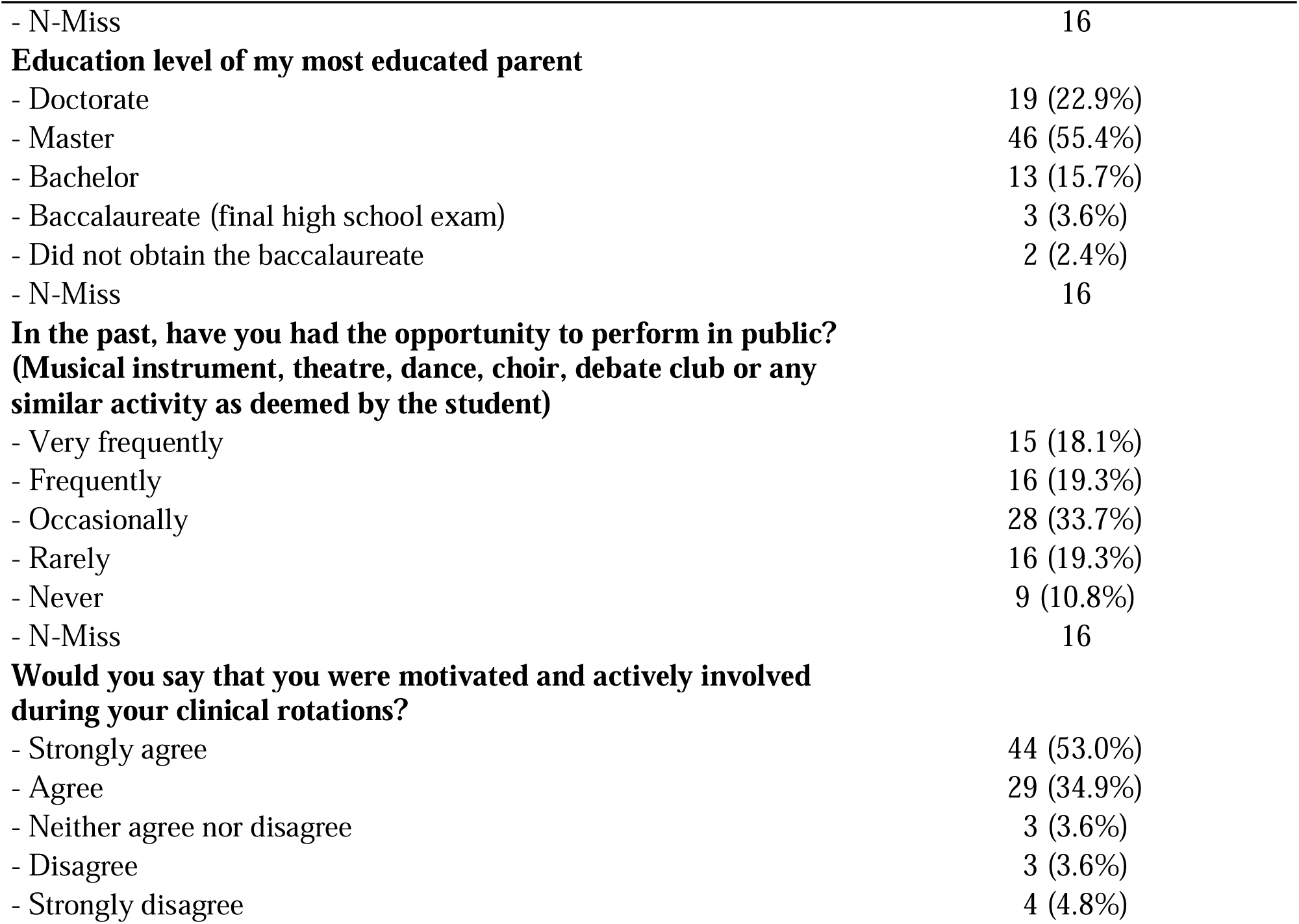
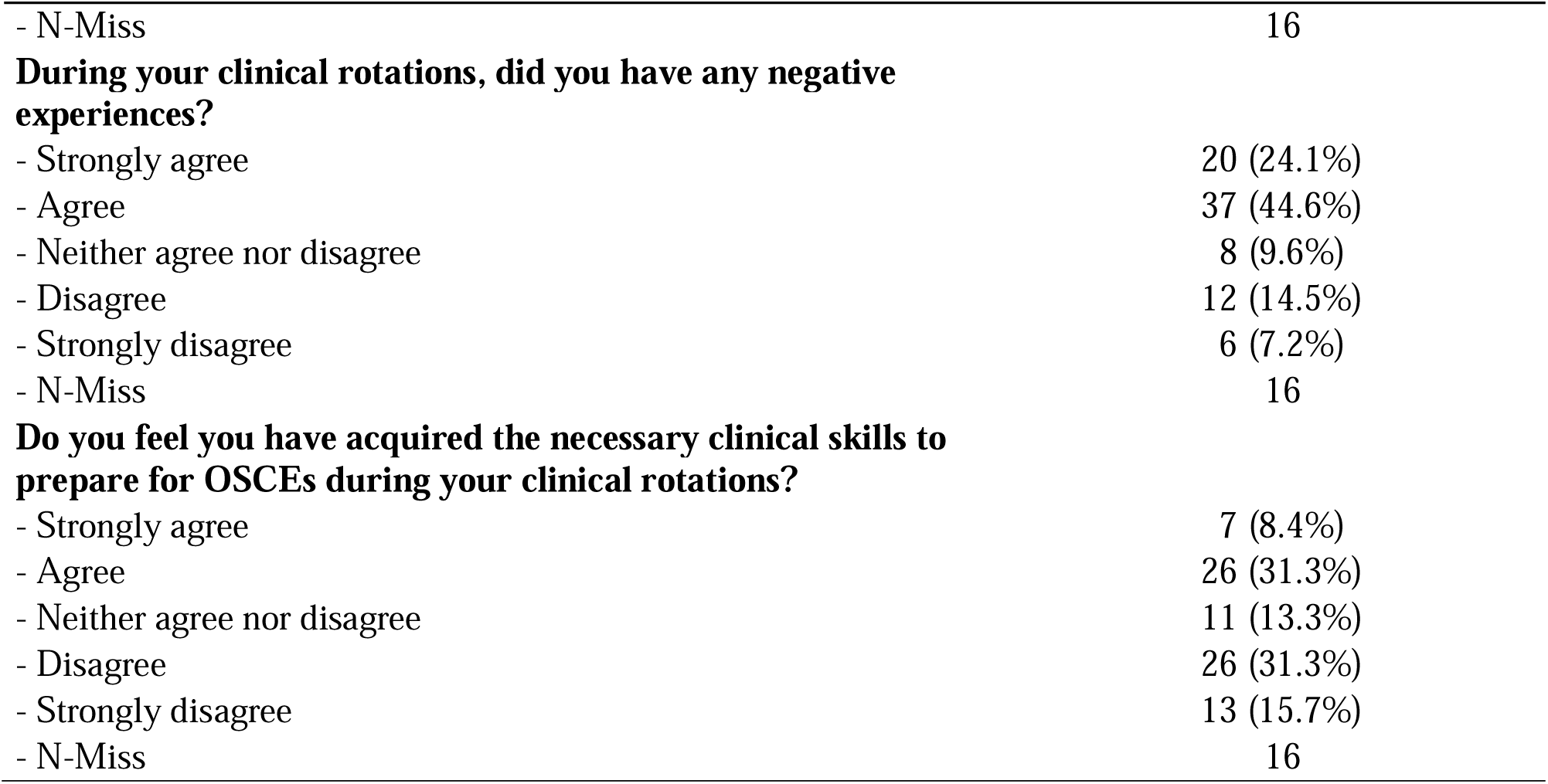
Participants’ Socio-Demographic Characteristics and Experiences During Clinical Rotations.

|  | Population (n=99) |
| --- | --- |
| **Gender** |  |
| - Male | 23 (27.7%) |
| - Female | 60 (72.3%) |
| - N-Miss | 16 |
| **Age** |  |
| - Mean (SD) | 23.04 (3.74) |
| - Median (Q1, Q3) | 22 (22, 23) |
| - N-Miss | 16 |
| **Future specialty** |  |
| - Medical | 55 (66.3%) |
| - Surgical | 23 (27.7%) |
| - Other | 5 (6.0%) |
| - N-Miss | 16 |
| **Parenthood** |  |
| - No child | 81 (97.6%) |
| - At least one child | 2 (2.4%) |
| - N-Miss | 16 |
| **Native language** |  |
| - French | 77 (92.8%) |
| - Other | 6 (7.2%) |
| - N-Miss | 16 |
| **Parents' native language** |  |
| - French for both | 59 (71.1%) |
| - The native language of at least one of my parents is not French | 24 (28.9%) |
| - N-Miss | 16 |
| **Education level of my most educated parent** |  |
| - Doctorate | 19 (22.9%) |
| - Master | 46 (55.4%) |
| - Bachelor | 13 (15.7%) |
| - Baccalaureate (final high school exam) | 3 (3.6%) |
| - Did not obtain the baccalaureate | 2 (2.4%) |
| - N-Miss | 16 |
| **In the past, have you had the opportunity to perform in public?**<br>**(Musical instrument, theatre, dance, choir, debate club or any**<br>**similar activity as deemed by the student)** |  |
| - Very frequently | 15 (18.1%) |
| - Frequently | 16 (19.3%) |
| - Occasionally | 28 (33.7%) |
| - Rarely | 16 (19.3%) |
| - Never | 9 (10.8%) |
| - N-Miss | 16 |
| **Would you say that you were motivated and actively involved**<br>**during your clinical rotations?** |  |
| - Strongly agree | 44 (53.0%) |
| - Agree | 29 (34.9%) |
| - Neither agree nor disagree | 3 (3.6%) |
| - Disagree | 3 (3.6%) |
| - Strongly disagree | 4 (4.8%) |
| - N-Miss | 16 |
| **During your clinical rotations, did you have any negative experiences?** |  |
| - Strongly agree | 20 (24.1%) |
| - Agree | 37 (44.6%) |
| - Neither agree nor disagree | 8 (9.6%) |
| - Disagree | 12 (14.5%) |
| - Strongly disagree | 6 (7.2%) |
| - N-Miss | 16 |
| **Do you feel you have acquired the necessary clinical skills to prepare for OSCEs during your clinical rotations?** |  |
| - Strongly agree | 7 (8.4%) |
| - Agree | 26 (31.3%) |
| - Neither agree nor disagree | 11 (13.3%) |
| - Disagree | 26 (31.3%) |
| - Strongly disagree | 13 (15.7%) |
| - N-Miss | 16 |

### Primary analysis

None of the four psychometric tests significantly correlated with OSCE scores.

We conducted univariate and multivariate analyses including the five domains of BFI-2, the three dimensions of the JSPE-MS, the SMM, and the GMS, the written mock examination scores and previous OSCE scores, **Table 2**. Conscientiousness significantly predicted OSCE scores in the univariate analysis (p=0.046), but not in the multivariate analysis (p=0.312). Performance in previous OSCE (December 13^th^, 2023) appeared to be the strongest positive predictor in both the univariate (p<0.001) and multivariate analyses (p=0.018), whereas performance in written mock examination was only a significant positive predictor in the univariate analysis (p=0.016). Being a male student showed a negative association in model (p-values = 0.051) and none of the four psychometric tests significantly predicted OSCE scores.

**Table 2:** Univariate and multivariate linear model explaining OSCE performance.

| Variable | Univariate analysis: $\beta$ (95% CI) | P-value | Multivariate analysis: $\beta$ (95% CI) | P-value |
| --- | --- | --- | --- | --- |
| **BFI2-Fr** |  |  |  |  |
| - Openness | -0.01 (-0.05 to 0.04) | 0.739 | 0 (-0.049 to 0.049) | 0.991 |
| - Conscientiousness | 0.05 (0.00 to 0.10) | 0.046** | 0.027 (-0.025 to 0.079) | 0.312 |
| - Extraversion | 0.01 (-0.03 to 0.05) | 0.779 | 0.002 (-0.043 to 0.046) | 0.934 |
| - Agreeableness | -0.01 (-0.07 to 0.05) | 0.761 | -0.021 (-0.097 to 0.054) | 0.575 |
| - Neuroticism | -0.00 (-0.04 to 0.03) | 0.859 | 0.03 (-0.017 to 0.077) | 0.209 |
| **JSPE-MS** |  |  |  |  |
| - Dimension 1 | -0.02 (-0.08 to 0.04) | 0.552 | 0.015 (-0.061 to 0.092) | 0.689 |
| - Dimension 2 | -0.06 (-0.14 to 0.02) | 0.119 | -0.063 (-0.162 to 0.036) | 0.206 |
| - Dimension 3 | -0.02 (-0.15 to 0.10) | 0.717 | -0.093 (-0.232 to 0.045) | 0.183 |
| - Total | -0.02 (-0.06 to 0.01) | 0.227 | NA |  |
| **Mindset scales** |  |  |  |  |
| - SMM | 0.03 (-0.01 to 0.06) | 0.126 | 0.022 (-0.012 to 0.055) | 0.198 |
| - GMS | -0.01 (-0.03 to 0.01) | 0.429 | -0.003 (-0.024 to 0.017) | 0.733 |
| - Total (sum) | 0.00 (-0.02 to 0.02) | 0.933 | NA |  |
| **Demographic characteristics** |  |  |  |  |
| - Age | -0.02 (-0.12 to 0.08) | 0.714 | 0.05 (-0.064 to 0.165) | 0.381 |
| - Sex (M as compared to F) | -0.75 (-1.66 to 0.16) | 0.106 | -1.136 (-2.276 to 0.003) | 0.051* |
| **Written mock examination** | 0.26 (0.05 to 0.48) | 0.016** | 0.19 (-0.056 to 0.437) | 0.128 |
| **Previous OSCE scores** | 0.31 (0.15 to 0.47) | <0.001*** | 0.234 (0.041 to 0.427) | 0.018** |

\*P≤0.05 (to two decimal places), \*\*P<0.050, \*\*\*P<0.010

**Table 3** presents the multivariate linear regression analyses assessing performance in interactive and non-interactive OSCE stations. Personality traits (BFI2-Fr) and demographic characteristics were included as predictors. In interactive OSCE stations (i.e., n°1,3,4), none of the personality traits or demographic variables significantly predicted performance. In non-interactive OSCE stations (i.e., n°2,5), Conscientiousness was the only significant predictor, with a positive association (p=0.001). Similar analyses were conducted with the other psychometric tests, and none of them yielded significant results.

**Table 3:** Multivariate linear models explaining performance in interactive and non-interactive OSCE stations.

| Variable | Linear models: $\beta$ (95% CI) | P-value | Linear models: $\beta$ (95% CI) | P-value |
| --- | --- | --- | --- | --- |
| **Interactive OSCE stations** |  |  |  |  |
| **BFI2-Fr** | **Multivariate analysis: $\beta$ (95% CI)** |  | **Multivariate analysis: $\beta$ (95% CI)** |  |
| - Openness | -0.104 (-0.257 to 0.049) | 0.180 | -0.080 (-0.235 to 0.076) | 0.312 |
| - Conscientiousness | 0.010 (-0.165 to 0.184) | 0.914 | -0.043 (-0.226 to 0.141) | 0.645 |
| - Extraversion | 0.003 (-0.144 to 0.149) | 0.971 | -0.040 (-0.193 to 0.114) | 0.608 |
| - Agreeableness | -0.053 (-0.262 to 0.156) | 0.614 | -0.083 (-0.313 to 0.148) | 0.479 |
| - Neuroticism | -0.009 (-0.149 to 0.131) | 0.893 | -0.070 (-0.227 to 0.087) | 0.378 |
| **Demographic characteristics** | **Univariate analysis: $\beta$ (95% CI)** |  | *Included in the multivariate analysis* |  |
| - Age | -0.118 (-0.455 to 0.218) | 0.486 | -0.046 (-0.424 to 0.333) | 0.811 |
| - Gender (M as compared to F) | -2.202 (-5.187 to 0.783) | 0.146 | -3.091 (-6.806 to 0.624) | 0.101 |
| **Non interactive OSCE stations** |  |  |  |  |
| **BFI2-Fr** | **Multivariate analysis: $\beta$ (95% CI)** |  | **Multivariate analysis: $\beta$ (95% CI)** |  |
| - Openness | 0.041 (-0.091 to 0.173) | 0.540 | 0.045 (-0.093 to 0.183) | 0.516 |
| - Conscientiousness | 0.272 (0.121 to 0.422) | 0.001*** | 0.270 (0.108 to 0.431) | 0.001*** |
| - Extraversion | 0.045 (-0.082 to 0.172) | 0.481 | 0.049 (-0.086 to 0.184) | 0.473 |
| - Agreeableness | -0.036 (-0.216 to 0.145) | 0.697 | -0.056 (-0.260 to 0.148) | 0.585 |
| - Neuroticism | 0.078 (-0.043 to 0.199) | 0.202 | 0.074 (-0.065 to 0.212) | 0.292 |
| **Demographic characteristics** | **Univariate analysis: $\beta$ (95% CI)** |  | *Included in the multivariate analysis* |  |
| - Age | 0.025 (-0.290 to 0.340) | 0.875 | 0.087 (-0.247 to 0.421) | 0.606 |
| - Gender (M as compared to F) | -1.516 (-4.317 to 1.286) | 0.285 | -0.205 (-3.484 to 3.074) | 0.901 |

\*P ≤ 0.100 (trends), \*\*P<0.050, \*\*\*P<0.010

We attempted to identify predictors of improvement in OSCE scores by comparing previous and studied OSCE results. Neuroticism showed a positive Spearman correlation coefficient of 0.224 (p=0.042). Also presented is a heatmap that includes the 15 facets of the BFI2-Fr and the five OSCE stations. The BFI2 facets of emotional lability and depression seem to be associated with lower scores, while assertiveness and responsibility appear to be associated with higher performance, **Supplementary material**.

### Secondary analyses

We further investigated the association between gender and these tests. Male students scored lower than female students in neuroticism (N) (OR=0.94, 95% CI [0.89-0.99], p=0.026), and empathy (OR=0.94, 95% CI [0.89-0.99], p=0.018). We investigated univariate analysis between OSCE scores and all demographic characteristics and commitment to clinical rotations. No significant correlations were found, **Supplementary material**.

The exploratory *post hoc* cluster analysis showed four distinct profiles. Cluster 4 (n=22) showed the highest Extraversion, Agreeableness, Conscientiousness, and Openness, the lowest Neuroticism, consistently high empathy across all three JSPE dimensions, a positive stress mindset, and a relatively high growth mindset. In contrast, Cluster 3 (n=21) showed high Neuroticism and Openness alongside strong emotional empathy (JSPE-MS dimension 3) but the lowest stress mindset. Cluster 2 (n=16) was characterised by high Agreeableness, strong cognitive empathy (JSPE-MS dimensions 1 and 2), and the highest Growth mindset, though with moderately elevated Neuroticism. Cluster 1 (n=25) showed lower empathy, lower Growth mindset, and less favorable Big Five traits, particularly Agreeableness and Openness. When examining the performance of these four student groups across the two OSCEs and the written examinations, cluster 4 consistently demonstrated the highest performance, whereas cluster 2 showed the lowest in two out of three examinations, **Supplementary material**.

## Discussion

We conducted a prospective cohort study, and contrary to initial hypotheses, neither personality domains, empathy nor stress-related mindsets were significantly associated with higher OSCE scores. To our knowledge, the 60-item BFI-2, GMS, and SMM have never been used in this context.

Nguyen *et al.* conducted a similar study [8]. They found no correlation between OSCE scores, empathy, and personality traits. Despite a non-significant trend, we were unable to produce conclusive data supporting the view that conscientiousness is an independent non-cognitive predictor of overall OSCE success. Although a recent comprehensive systematic review by Chisholm-Burns *et al.* reports that conscientiousness is associated with “clinical/experiential performance,” this broader category does not correspond specifically to OSCEs [20]. Other recent studies do not find any correlation between conscientiousness (C) and OSCE scores [5,7]. Neuroticism and extraversion have also been linked to higher OSCE scores [6].

Self-reported empathy was not associated with higher scores, as reported by Casas *et al.* [9]. Other studies, could not establish a correlation with higher OSCE scores [7–10]. Intuitively, higher stress and anxiety levels are negative predictive factors. Many studies do not find correlations with OSCE scores [11–15]. Nonetheless, one study found a negative correlation [46] and another indicates that a stress management intervention can improve OSCE scores [16]. This finding is consistent with a recent systematic review by Martin *et al.* [15], although most studies included used the State-Trait Anxiety Inventory [11,47–49], and none used mindset scales.

Although the non-cognitive factors investigated here do not correlate with higher OSCE scores, this does not mean that OSCEs are irrelevant to the training of medical students. To implement pertinent and innovative pedagogical strategies, identifying the factors that can be changed over time is necessary. Abbiati *et al.* evaluated the Big Five personality domains of 244 medical students in their 1^st^ and 6^th^ years of training [6]. They concluded that personality remains constant throughout medical school. Thus, interventions aiming at modifying personality may not be relevant. Empathy, however, shows greater variability. A meta-analysis by Spatoula *et al.* shows a decrease during undergraduate training [50]. Nonetheless, a systematic review by Andersen *et al.* presents more nuanced results and cautions against hasty conclusions [51]. Specifically, empathy as measured with the JSPE may only capture the value students place upon clinical empathy rather than effective empathy skills [52].

If these non-cognitive factors can change over time, it is necessary to identify feasible, relevant, and cost-effective interventions. For instance, Balint group training consisting of seven sessions of an hour and a half spread over two months has been found to improve medical students’ empathy [53]. A systematic review by Kelm *et al.* provides insight into the various interventions linked to an increase in empathy [54]. Concerning stress-related mindsets, short online interventions can substantially change students’ mindsets [37,55]. Furthermore, potential interventions based on medical humanities could be explored [56,57].

The finding that male gender is associated with lower OSCE performance has been reported in a meta-analysis by Woolf et al. [58], while a critical literature review by Chao et al. did not identify such an association [59].

Conscientiousness was a significant predictor of performance in non-interactive OSCE stations. This finding suggests that OSCE results should not be considered a homogeneous entity. The association between academic success and conscientiousness has been frequently reported in the literature [20,60–62], and it could be hypothesised that non-interactive stations resemble a traditional university oral examination.

The doctor-patient relationship is highly complex, involving multiple cognitive and non-cognitive skills. OSCEs are a highly pertinent modality for assessing clinical skills but may not be as reliable as expected in encompassing all the required qualities, especially empathy. The results of our study should not be taken as undermining the value of OSCEs. On the contrary, it should call for innovation in pedagogy and further research, such as implementing complementary assessments (e.g., situational judgment tests, structured self-reflection exercises) to capture these dimensions, or developing more comprehensive assessment tools.

OSCEs evaluate the application and integration of knowledge in clinical settings, aligning with the higher levels of Bloom’s taxonomy (application, analysis, evaluation) [63]. In contrast, psychometric scales primarily rely on self-perception and self-reported attitudes, raising concerns about their ecological validity. Self-assessments are often influenced by social desirability bias, a lack of self-awareness, or the absence of real-life situational context, limiting their ability to predict actual performance. For example, students may perceive themselves as empathetic without effectively demonstrating empathy in a real clinical interaction. While OSCEs provide direct observation of a student’s performance in a standardized clinical scenario, psychometric scales offer insights into students’ predispositions and self-perceived competencies. A comprehensive assessment of non-cognitive skills should integrate multiple approaches, combining direct observation (OSCEs), real or simulated patient feedback, and self-assessments to capture both observed behaviors and underlying dispositions.

### Limitations

One of the limitation of our study is its observational design, which results in an unavoidable selection bias. To assess this difficulty, we compared our cohort with the overall fifth-year medical students’ cohort. It appears that both cohorts had similar OSCEs scores and gender representation. However, the participants in our study were slightly younger and achieved higher scores on the written mock examination and previous OSCEs, similar to findings reported by Nguyen *et al.*[7]. When participation is voluntary, this bias seems difficult to prevent.

Another limitation is the monocentric design. However, each university administers its own OSCEs and assesses them on different dates.

In addition, the participation rate for the questionnaire was low. Nevertheless, the number of participants is comparable to that of several studies on similar topics [8,10–12,14,16,24].

Finally, as OSCEs are a more subjective assessment tool than written examinations, the absence of correlation between potential predictors and OSCE results could be related to the data’s lack of robustness. Personality domains, empathy, and stress-related mindsets were self-reported and therefore subjective. Ogle *et al.* proposed that empathy [8] could have been assessed by simulated patients or a neutral observer. Moreover, concerns have been raised about the intrinsic reliability of OSCEs [64], especially for assessing communication skills [65]. Several tools have been suggested to investigate and tackle this issue [66].

## Conclusion

OSCEs are a validated and widely used method for assessing clinical competence in medical education. However, student performance could not be reliably predicted based on personality traits, self-reported empathy, or stress-related mindsets. Conscientiousness emerged as a significant predictor of performance in non-interactive OSCE stations, while neuroticism was positively associated with performance improvement across OSCE sessions. Multidimensional psychometric assessment may be relevant when investigating performance outcomes in OSCEs. Further studies are warranted to explore how self-assessment tools could be integrated into OSCEs to better capture the full spectrum of non-cognitive skills among medical students.

## Data Availability

The datasets used during this study are available from the corresponding author on reasonable request.

## Declarations

### Consent for publication

Not applicable.

### Consent to participate

Not applicable.

### Statement on the Use of Artificial Intelligence

Not applicable.

### Authors’ contributions

Conception and design: DH, AG. Acquisition of data: DH. Analysis and interpretation of data: DH, BL, BM, OP, Université Paris Cité OSCEs study group, CL, AG. Drafting the article: DH, AG. Critical revision of the article: BL, BM, OP, Université Paris Cité OSCEs study group, CBDV, DB, AF, CL. Review of submitted version of manuscript: DH, AG. Approval of the final version of the manuscript on behalf of all authors: AG.

### Ethical approval

The Institutional Review Board (IRB) of the Université Paris Cité authorised our study (number 00012024-21). Informed consent was obtained from all participants.

## Acknowledgements

We would like to thank Dr. Cédric Thépenier from the Institut Pasteur, all the Université Paris Cité staff, volunteers, and the Université Paris Cité OSCEs study group: Albert Faye, Donia Bouzid, Ugo Marchese, Michael Thy, Anna Pellat, Gilles Soulat, Alexy Tran Dinh, Valentine Ferré, Nathan Peiffer Smadja, Benjamin Deniau, Bénédicte Oules, Yann Nguyen, Lina Khider, Thibaud Soumagne, Augustin Gaudemer, Idir Ouzaid, Sophie Mazar, Jean Marc Liger, Eric Desrentes, Léonore Muller.

The authors wish to thank the Thomas Jefferson University Medical College for permission to use the Jefferson Scale of Physician Empathy-Medical Student, Dr Alexandre Malmartel, lecturer at Université Paris Cité, who published the French-validated translation of the JSPE-MS, the Department of Psychology of Stanford University for their permission to use the Stress Mindset Measure and the Growth Mindset Scale, the Centre Laennec Paris, TEAM UP student tutoring association and students representatives at Université Paris Cité for helping us promote our study among fifth-year medical students.

**Supplementary material:** 1) Description of the five OSCE stations, 2) Questionnaire, 3) Comparison between the study cohort and the overall fifth-year medical students’ cohort, 4) Spearman correlation coefficients between the change in OSCE scores across two sessions and the psychometric tests, 5) Heatmap, 6) Univariate linear model explaining OSCE scores by demographic characteristics, 7) Exploratory cluster analysis

