## Supplementary material for "Beyond skills: The impact of personality traits, empathy and stress mindset on OSCE outcomes"

**Table** Description of the five OSCE stations

| **Station number** | **Station type** | **Human interaction** | **Primary Tasks** |
| --- | --- | --- | --- |
| **1** | Standardized Patient | Yes | Taking a medical history, analyzing documentation, and deciding on hospital admission |
| **2** | Without a standardized patient or healthcare professional | No | Analyzing clinical data, establishing a diagnosis, formulating a therapeutic strategy, and oral reasoning |
| **3** | Standardized Patient | Yes | Conducting a medical history interview, explaining the diagnosis, and ordering tests |
| **4** | Standardized Healthcare Professional | Yes | Engaging in interprofessional communication, explaining the diagnosis, and making therapeutic proposals |
| **5** | Procedural | No | Breast palpation (including explanation and performance), clinical conclusion, and suggesting tests |

*OSCEs include various types of stations: with a standardised patient, with a standardised healthcare professional, without a standardised patient or healthcare professional, and procedural stations. The scoring grid consists of 9 to 12 binary items (0 or 1), mainly focusing on medical knowledge, 2 to 5 items assessing communication and attitudes skills (on a 5-point Likert scale), and an overall performance item (on a 5-point Likert scale). Items scored on a Likert scale, such as non-verbal communication, ability to listen, and ability to cooperate with peers, seem to be more likely related to non-cognitive skills.*

**Questionnaire**

**Pedagogical Study**

**Personality Assessment: Big Five Inventory - 2 Fr**
You will find a series of descriptive statements that may or may not apply to you. For example, are you someone who enjoys spending time with others?
After each statement, select the degree to which you agree or disagree with it.

**I am someone who...**

*****

Choose the appropriate answer for each item:

|  | **Disagree strongly** | **Disagree a little** | **Neutral; no opinion** | **Agree a little** | **Agree strongly** |
| --- | --- | --- | --- | --- | --- |
| **Is outgoing, sociable** |  |  |  |  |  |
| **Is compassionate, has a soft heart** |  |  |  |  |  |
| **Tends to be disorganized** |  |  |  |  |  |
| **Is relaxed, handles stress well** |  |  |  |  |  |
| **Has few artistic interests** |  |  |  |  |  |
| **Has an assertive personality** |  |  |  |  |  |
| **Is respectful, treats others with respect** |  |  |  |  |  |
| **Tends to be lazy** |  |  |  |  |  |
| **Stays optimistic after experiencing a setback** |  |  |  |  |  |
| **Is curious about many different things** |  |  |  |  |  |
| **Rarely feels excited or eager** |  |  |  |  |  |
| **Tends to find fault with others** |  |  |  |  |  |
| **Is dependable, steady** |  |  |  |  |  |
| **Is moody, has up and down mood swings** |  |  |  |  |  |
| **Is inventive, finds clever ways to do things** |  |  |  |  |  |

*****

Select the appropriate response for each item:

|  | **Disagree strongly** | **Disagree a little** | **Neutral; no opinion** | **Agree a little** | **Agree strongly** |
| --- | --- | --- | --- | --- | --- |
| **Tends to be quiet** |  |  |  |  |  |
| **Feels little sympathy for others** |  |  |  |  |  |
| **Is systematic, likes to keep things in order** |  |  |  |  |  |
| **Can be tense** |  |  |  |  |  |
| **Is fascinated by art, music, or literature** |  |  |  |  |  |
| **Is dominant, acts as a leader** |  |  |  |  |  |
| **Starts arguments with others** |  |  |  |  |  |
| **Has difficulty getting started on tasks** |  |  |  |  |  |
| **Feels secure, comfortable with self** |  |  |  |  |  |
| **Avoids intellectual, philosophical discussions** |  |  |  |  |  |
| **Is less active than other people** |  |  |  |  |  |
| **Has a forgiving nature** |  |  |  |  |  |
| **Can be somewhat careless** |  |  |  |  |  |
| **Is emotionally stable, not easily upset** |  |  |  |  |  |
| **Has little creativity** |  |  |  |  |  |

*****

Select the appropriate response for each item:

|  | **Disagree strongly** | **Disagree a little** | **Neutral; no opinion** | **Agree a little** | **Agree strongly** |
| --- | --- | --- | --- | --- | --- |
| **Is sometimes shy, introverted** |  |  |  |  |  |
| **Is helpful and unselfish with others** |  |  |  |  |  |
| **Keeps things neat and tidy** |  |  |  |  |  |
| **Worries a lot** |  |  |  |  |  |
| **Values art and beauty** |  |  |  |  |  |
| **Finds it hard to influence people** |  |  |  |  |  |
| **Is sometimes rude to others** |  |  |  |  |  |
| **Is efficient, gets things done** |  |  |  |  |  |
| **Often feels sad** |  |  |  |  |  |
| **Is complex, a deep thinker** |  |  |  |  |  |
| **Is full of energy** |  |  |  |  |  |
| **Is suspicious of others’ intentions** |  |  |  |  |  |
| **Is reliable, can always be counted on** |  |  |  |  |  |
| **Keeps their emotions under control** |  |  |  |  |  |
| **Has difficulty imagining things** |  |  |  |  |  |

*****

Select the appropriate response for each item:

|  | **Disagree strongly** | **Disagree a little** | **Neutral; no opinion** | **Agree a little** | **Agree strongly** |
| --- | --- | --- | --- | --- | --- |
| **Is talkative** |  |  |  |  |  |
| **Can be cold and uncaring** |  |  |  |  |  |
| **Leaves a mess, doesn’t clean up** |  |  |  |  |  |
| **Rarely feels anxious or afraid** |  |  |  |  |  |
| **Thinks poetry and plays are boring** |  |  |  |  |  |
| **Prefers to have others take charge** |  |  |  |  |  |
| **Is polite, courteous to others** |  |  |  |  |  |
| **Is persistent, works until the task is finished** |  |  |  |  |  |
| **Tends to feel depressed, blue** |  |  |  |  |  |
| **Has little interest in abstract Ideas** |  |  |  |  |  |
| **Shows a lot of Enthusiasm** |  |  |  |  |  |
| **Assumes the best about people** |  |  |  |  |  |
| **Sometimes behaves irresponsibly** |  |  |  |  |  |
| **Is temperamental, gets emotional easily** |  |  |  |  |  |
| **Is original, comes up with new Ideas** |  |  |  |  |  |

**Empathy Assessment: JSPE-MS© French Version**

Indicate the degree to which you agree or disagree with each statement by selecting the appropriate number from 1 (strongly disagree) to 7 (strongly agree).

© Thomas Jefferson University, 2001 All rights reserved

*****

*The JSPE-MS© is subject to copyright and cannot be fully reproduced without permission.*

**Assessment of Stress Perception: Stress Mindset Measure & Growth Mindset Scale**
Indicate your level of agreement or disagreement with each of the following statements.

*****

Select the appropriate response for each item:

|  | Strongly Disagree | Disagree | Neither Agree nor Disagree | Agree | Strongly Agree |
| --- | --- | --- | --- | --- | --- |
| 1. The effects of stress are negative and should be avoided.* |  |  |  |  |  |
| 2. Experiencing stress facilitates my learning and growth. |  |  |  |  |  |
| 3. Experiencing stress depletes my health and vitality.* |  |  |  |  |  |
| 4. Experiencing stress enhances my performance and productivity. |  |  |  |  |  |
| 5. Experiencing stress inhibits my learning and growth.* |  |  |  |  |  |
| 6. Experiencing stress improves my health and vitality. |  |  |  |  |  |
| 7. Experiencing stress debilitates my performance and productivity.* |  |  |  |  |  |
| 8. The effects of stress are positive and should be utilized. |  |  |  |  |  |

*****

Select the appropriate response for each item:

|  | strongly disagree | disagree | mostly disagree | mostly agree | agree | strongly agree |
| --- | --- | --- | --- | --- | --- | --- |
| You have a certain amount of intelligence, and you can’t really do much to change it. |  |  |  |  |  |  |
| Your intelligence is something about you that you can’t change very much. |  |  |  |  |  |  |
| You can learn new things, but you can’t really change your basic intelligence. |  |  |  |  |  |  |

**Clinical Rotation and General Demographic Characteristics**

**Indicate your level of agreement or disagreement with each of the following statements. ***
Select the appropriate response for each item:

|  | Strongly disapprove | Neither approve nor disapprove | Neither approve nor disapprove | Somewhat approve | Strongly approve |
| --- | --- | --- | --- | --- | --- |
| Would you say that you were motivated and actively involved during your clinical rotations? |  |  |  |  |  |
| During your clinical rotations, did you experience any negative events? |  |  |  |  |  |
| Do you feel you acquired the clinical skills necessary to prepare for OSCEs during your clinical rotations? |  |  |  |  |  |

**Gender ***

Please select only one of the following options:

- Male
- Female
- Other

**Age ***

Your answer must be between 0 and 99.
Please write your answer here:

**Intended specialty ***

Please select only one of the following options:

- Medical
- Surgical
- Other

**Parenthood (Are you a parent?) ***

Please select only one of the following options:

- No children
- At least one child

**Native language***

Please select only one of the following options:

- French
- Other

**Parents' native language** *****

Please select only one of the following options:

- French for both
- At least one of my parents' native language is not French

**Education level of my most educated parent** *****

Please select only one of the following options:

- Did not obtain the Baccalaureate
- Baccalaureate
- Bachelor's degree
- Master's degree
- Doctorate

**In the past, have you had the opportunity to perform in public? (e.g., playing a musical instrument, theater, dance, choir, debate club, or any other similar activity as deemed relevant by the student)***

Choose the appropriate answer for each item:

|  | **Never** | **Rarely** | **Sometimes** | **Often** | **Very often** |
| --- | --- | --- | --- | --- | --- |

**Comparison between the study cohort and the overall fifth-year medical students’ cohort**

| **Test** | **label** | **levels** | **Study cohort** | **Overall cohort** | **Uncorrected p-values** | **Corrected p-values** |
| --- | --- | --- | --- | --- | --- | --- |
| **Previous OSCE** | s01 | Median (IQR) | 17.1 (13.7 to 19.8) | 15.7 (12.7 to 18.3) | 0.066 | 0.215 |
|  | s02 | Median (IQR) | 12.6 (9.7 to 17.2) | 11.5 (8.6 to 16.4) | 0.250 | 0.343 |
|  | s03 | Median (IQR) | 13.4 (9.5 to 17.9) | 12.5 (9.6 to 15.7) | 0.085 | 0.221 |
|  | s04 | Median (IQR) | 15.3 (13.3 to 17.7) | 14.6 (11.8 to 17.1) | 0.026 | 0.113 |
|  | s05 | Median (IQR) | 14.4 (12.8 to 16.5) | 14.1 (11.1 to 16.5) | 0.264 | 0.343 |
|  | TOT | Median (IQR) | 14.5 (12.9 to 16.0) | 13.7 (12.0 to 15.1) | 0.009** | 0.065 |
| **Studied OSCE** | s01 | Median (IQR) | 15.4 (13.0 to 17.5) | 15.1 (12.8 to 17.2) | 0.244 | 0.343 |
|  | s02 | Median (IQR) | 16.6 (14.1 to 19.6) | 16.6 (12.3 to 19.9) | 0.622 | 0.622 |
|  | s03 | Median (IQR) | 13.4 (11.5 to 15.3) | 13.3 (11.3 to 15.1) | 0.330 | 0.39 |
|  | s04 | Median (IQR) | 12.7 (10.9 to 14.3) | 13.0 (10.7 to 14.9) | 0.500 | 0.542 |
|  | s05 | Median (IQR) | 14.1 (11.9 to 15.7) | 13.5 (11.6 to 15.2) | 0.158 | 0.342 |
|  | TOT | Median (IQR) | 14.2 (12.8 to 15.7) | 13.9 (12.6 to 15.3) | 0.195 | 0.343 |
| **Written mock examination** | TOT | Median (IQR) | 13.9 (12.9 to 14.7) | 13.4 (12.4 to 14.5) | 0.010* | 0.065 |
|  | Age | Median (IQR) | 22.0 (22.0 to 23.0) | 23.0 (22.0 to 24.0) | 0.008** | NA |
|  | Gender | Proportions | 76.6%(F), 23.4% | 67.8%(F), 32.2% | 0.124 | NA |

**Spearman correlation coefficients between the change in OSCE scores across two sessions and the psychometric tests**

| **Tests** | **Progression (OSCE – Previous OSCE)** |
| --- | --- |
| **bfi_Openness** | -0.039 (p: 0.728) |
| **bfi_Conscienciousness** | -0.059 (p: 0.598) |
| **bfi_Extraversion** | -0.109 (p: 0.326) |
| **bfi_Agreeableness** | 0.023 (p: 0.839) |
| **bfi_Neuriticism** | 0.224 (p: 0.042)* |
| **JSPE_dim1** | 0.027 (p: 0.809) |
| **JSPE_dim2** | 0.011 (p: 0.923) |
| **JSPE_dim3** | -0.073 (p: 0.511) |
| **JSPE_total** | 0.03 (p: 0.786) |
| **SMM** | -0.039 (p: 0.726) |
| **GMS** | 0.056 (p: 0.612) |
| **Mindsets** | 0.029 (p: 0.796) |

**Heatmap representing the Spearman correlation coefficients between the written mock examination, previous and studied OSCE stations, and the various psychometric tests including the 15 facets of the BFI2**

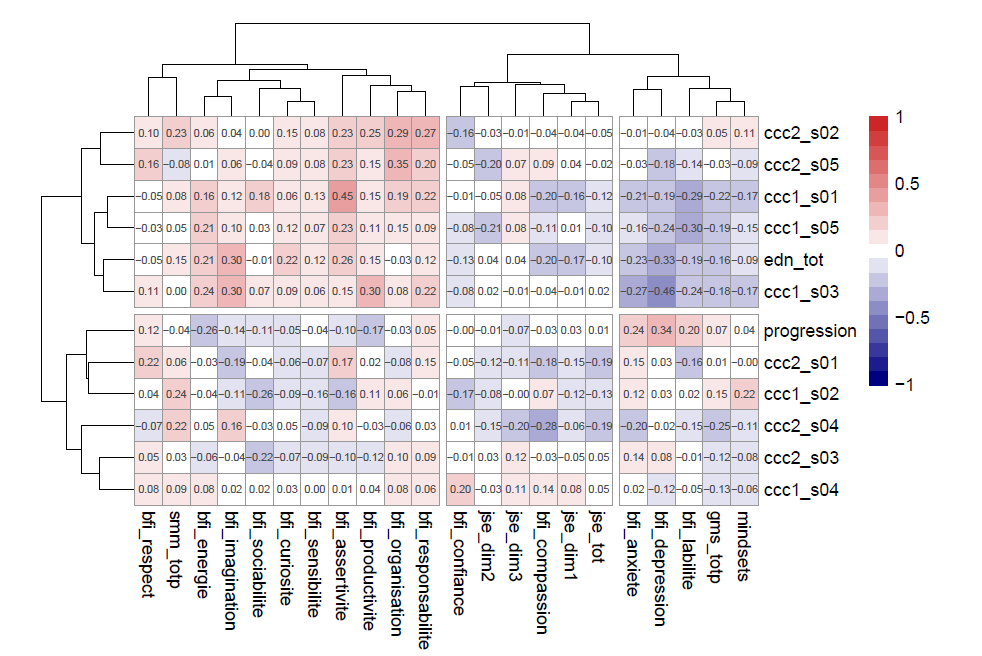

**Univariate linear model explaining OSCE scores by demographic characteristics**

| **Variable** | **Levels** | **Number** | **Median OSCE score (IQR)** | **Coefficient (univariable)** |
| --- | --- | --- | --- | --- |
| **age** | [2.0,40.0] | 77 (100.0) | 14.3 (13.4 to 15.7) | -0.02 (-0.12 to 0.08, p=0.714) |
| **gender** | F | 59 (76.6) | 14.3 (13.6 to 15.7) | - |
|  | M | 18 (23.4) | 13.9 (12.3 to 15.1) | -0.75 (-1.66 to 0.16, p=0.106) |
| **Intended specialty** | Medical | 49 (63.6) | 14.2 (13.5 to 15.5) | - |
|  | Surgical | 23 (29.9) | 14.4 (12.9 to 15.8) | 0.10 (-0.76 to 0.97, p=0.816) |
|  | Other | 5 (6.5) | 14.8 (14.7 to 16.4) | 1.22 (-0.39 to 2.83, p=0.135) |
| **Parenthood** | No children | 75 (97.4) | 14.3 (13.4 to 15.7) | - |
|  | At least one child | 2 (2.6) | 12.8 (11.8 to 13.7) | -1.56 (-4.01 to 0.89, p=0.208) |
| **Native language** | French | 71 (92.2) | 14.3 (13.4 to 15.7) | - |
|  | Other | 6 (7.8) | 14.1 (12.5 to 14.3) | -0.78 (-2.24 to 0.68, p=0.290) |
| **Parents' native language** | French for both | 54 (70.1) | 14.7 (13.5 to 15.8) | - |
|  | Other for ≥ 1 parent | 23 (29.9) | 14.1 (13.4 to 15.2) | -0.30 (-1.16 to 0.55, p=0.484) |
| **Education level of my most educated parent** | Did not obtain the Baccalaureate | 2 (2.6) | 14.4 (14.0 to 14.9) | - |
|  | Baccalaureate | 2 (2.6) | 16.8 (16.6 to 17.0) | 2.35 (-0.96 to 5.66, p=0.161) |
|  | Bachelor's degree | 13 (16.9) | 13.8 (12.3 to 14.9) | -1.06 (-3.57 to 1.45, p=0.404) |
|  | Master's degree | 42 (54.5) | 14.7 (13.6 to 15.8) | 0.09 (-2.31 to 2.48, p=0.943) |
|  | Doctorate | 18 (23.4) | 14.1 (13.1 to 14.8) | -0.47 (-2.94 to 1.99, p=0.704) |
| **In the past, have you had the opportunity to perform in public? (e.g., playing a musical instrument, theater, dance, choir, debate club, or any other similar activity as deemed relevant by the student)** | Never | 9 (11.7) | 13.5 (11.8 to 15.7) | - |
|  | Rarely | 13 (16.9) | 14.2 (13.4 to 15.8) | 0.56 (-0.95 to 2.08, p=0.460) |
|  | Sometimes | 26 (33.8) | 14.5 (12.9 to 15.7) | 0.40 (-0.95 to 1.75, p=0.555) |
|  | Often | 15 (19.5) | 14.3 (13.9 to 15.3) | 0.83 (-0.64 to 2.30, p=0.265) |
|  | Very often | 14 (18.2) | 14.4 (13.8 to 15.6) | 0.73 (-0.76 to 2.22, p=0.334) |
| **Would you say that you were motivated and actively involved during your clinical rotations?** | Strongly disapprove | 3 (3.9) | 11.9 (11.7 to 13.8) | - |
|  | Somewhat disapprove | 3 (3.9) | 16.0 (13.8 to 16.7) | 2.00 (-0.80 to 4.80, p=0.159) |
|  | Somewhat approve | 27 (35.5) | 14.7 (13.7 to 15.8) | 1.46 (-0.63 to 3.55, p=0.167) |
|  | Strongly approve | 43 (56.6) | 14.2 (13.4 to 15.7) | 1.17 (-0.88 to 3.22, p=0.259) |
| **During your clinical rotations, did you experience any negative events?** | Strongly disapprove | 6 (7.8) | 15.7 (14.3 to 15.8) | - |
|  | Somewhat disapprove | 10 (13.0) | 14.0 (13.7 to 14.4) | -1.23 (-3.02 to 0.56, p=0.174) |
|  | Neither approve nor disapprove | 7 (9.1) | 13.6 (12.9 to 14.2) | -1.22 (-3.15 to 0.70, p=0.210) |
|  | Somewhat approve | 35 (45.5) | 14.3 (13.6 to 15.2) | -0.65 (-2.18 to 0.88, p=0.397) |
|  | Strongly approve | 19 (24.7) | 15.7 (12.3 to 16.0) | -0.46 (-2.08 to 1.16, p=0.573) |
| **Do you feel you acquired the clinical skills necessary to prepare for OSCEs during your clinical rotations?** | Strongly disapprove | 11 (14.3) | 14.7 (12.1 to 15.8) | - |
|  | Somewhat disapprove | 25 (32.5) | 14.4 (13.6 to 15.7) | 0.21 (-1.06 to 1.47, p=0.747) |
|  | Neither approve nor disapprove | 10 (13.0) | 14.3 (14.1 to 14.7) | -0.29 (-1.82 to 1.23, p=0.702) |
|  | Somewhat approve | 25 (32.5) | 14.0 (13.4 to 15.7) | 0.09 (-1.17 to 1.36, p=0.883) |
|  | Strongly approve | 6 (7.8) | 12.8 (12.0 to 15.6) | -0.57 (-2.35 to 1.20, p=0.523) |
| **Written mock examination** | [1.8,16.8] | 77 (100.0) | 14.3 (13.4 to 15.7) | 0.26 (0.05 to 0.48, p=0.016)* |

|  | **Cluster 1 (N=25)** | **Cluster 2 (N=16)** | **Cluster 3 (N=21)** | **Cluster 4 (N=22)** | **p value** |
| --- | --- | --- | --- | --- | --- |
| **Extraversion** | 34.440 (9.051) | 31.000 (6.470) | 39.810 (8.830) | 46.182 (7.719) | < 0.001** |
| **Agreeableness** | 41.880 (6.254) | 49.625 (4.856) | 46.238 (5.890) | 51.955 (2.681) | < 0.001** |
| **Conscienciousness** | 45.880 (7.541) | 42.375 (7.822) | 42.810 (8.370) | 49.500 (6.624) | 0.010** |
| **Neuroticism** | 33.960 (9.414) | 40.750 (8.315) | 45.286 (10.090) | 30.273 (7.567) | < 0.001** |
| **Openness** | 39.360 (8.878) | 38.688 (7.171) | 49.143 (7.045) | 49.773 (5.806) | < 0.001** |
| **JSPE MS dimension 1** | 51.640 (5.559) | 58.688 (4.094) | 58.238 (5.440) | 59.364 (6.276) | < 0.001** |
| **JSPE MS dimension 2** | 40.440 (6.653) | 48.750 (3.256) | 51.190 (2.657) | 50.273 (2.434) | < 0.001** |
| **JSPE MS dimension 3** | 6.280 (2.208) | 4.562 (2.190) | 9.571 (2.874) | 7.636 (2.804) | < 0.001** |
| **SMM** | 13.120 (4.893) | 12.938 (5.916) | 7.286 (4.981) | 16.727 (6.504) | < 0.001** |
| **GMS** | 10.040 (2.791) | 13.625 (2.802) | 10.857 (3.941) | 11.818 (3.737) | 0.010** |

**Clustering of Participants Into Four Psychometric Profiles Using k-Means on Four Principal Components**

**P<0.050, ***P<0.010

**Comparison of OSCE and Written Examination Performance Across Four Groups**

|  | **Cluster 1 (N=25)** | **Cluster 2 (N=16)** | **Cluster 3 (N=21)** | **Cluster 4 (N=22)** | **p value** |
| --- | --- | --- | --- | --- | --- |
| **Age** | 22.375 (4.853) | 24.062 (4.582) | 23.571 (3.385) | 22.500 (1.058) | 0.834 |
| **Being Female** | 14 (58.3%) | 10 (62.5%) | 19 (90.5%) | 17 (77.3%) | 0.077* |
| **OSCE 1** | 14.528 (2.455) | 12.881 (2.352) ↓ | 14.433 (2.234) | 14.768 (1.824) ↑ | 0.087* |
| **OSCE 2** | 14.196 (2.021) | 13.794 (2.120) | 13.776 (1.742) ↓ | 14.468 (1.303) ↑ | 0.660 |
| **Written examination** | 13.790 (2.849) | 13.015 (1.058) ↓ | 13.865 (0.873) | 14.193 (1.254) ↑ | 0.009*** |

*P≤0.100 (trends), **P<0.050, ***P<0.010

↑ : highest score

↓ : lowest score

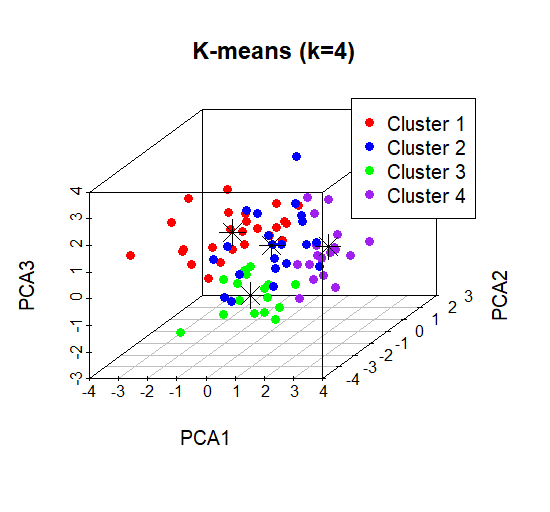

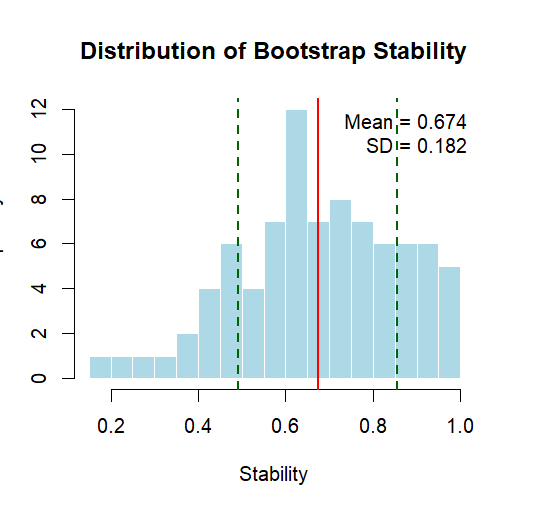
